# Quantification of Patent Foramen Ovale Shunt Severity by Transesophageal Echocardiogram and Transcranial Doppler in Routine Clinical Practice

**DOI:** 10.1101/2024.07.12.24310355

**Authors:** Philip Y. Sun, Jonathan M. Tobis, Samuel A. Daneshvar, Rodel C. Alfonso, David S. Liebeskind, Jeffrey L. Saver

## Abstract

**Background:** The presence of a large physiologic shunt, defined as >20 left atrial microbubbles within 3 cardiac cycles on transesophageal echocardiography (TEE) is a randomized controlled trial-validated indication for patent foramen ovale (PFO) closure in patients with otherwise cryptogenic ischemic stroke. The frequency with which this information is available to treating physicians from clinical TEE reports has not been well delineated.

**Methods:** Among consecutive ischemic stroke patients admitted to an academic medical center undergoing TEE between October 2022 - September 2023, TEE reports were reviewed to determine frequency with which formal clinical-trial definition-adherent characterization of PFO shunt size was provided, the frequency of informal PFO shunt size descriptions, and correlations with transcranial Doppler (TCD) formal shunt grades (Spencer Grades 1-5).

**Results:** Among 77 patients, the median age was 64 (IQR 56-73), and 33 (43%) were female. On TEE, shunt presence was assessed by bubble study in 60 (78%), direct Doppler alone in 5 (7%), and neither in 12 (16%). Among patients with bubble study, a right-to-left shunt (RLS) potentially due to PFO was present 25 (42%). RLS severity was quantified on the clinical report in 16% (4/25) of PFO patients, all with small (<20) shunts. For the remaining 21 (84%), reports informally characterized shunt severity with various descriptive terms, including “small/mild/trace” (13 cases), “moderate/medium” (6), and “large” (1). TCD bubble studies were performed in 76% (19/25) of the TEE RLS PFO patients. Shunt severity correlation between TEE and TCD was 100% (3/3) for the formally quantified TEE shunts, un- assessable in one patient with non-specific shunt size characterization on TEE, and poor (3/15, 20%) for the remaining 15 informally assessed TEE shunts, with 47% (7/15) of shunts categorized as larger on TCD and 33% (5/15) smaller on TCD than TEE.

**Conclusions:** Quantified, evidence-based ratings of PFO shunt severity were present in less than 1 of every 6 TEE reports, and unquantified, informal size estimates correlated poorly with TCD quantification of shunt severity. Patient management would be aided by inclusion of formal PFO shunt size quantification in all clinical stroke patient TEE reports.

## Introduction

Stroke by paradoxical embolism through a patent foramen ovale (PFO), a structural variant present in about a quarter of adults, is a common cause of ischemic stroke, accounting for 5% of all cases. However, PFOs also frequently are incidentally present and not causally related to an otherwise cryptogenic stroke^1,2^. Risk stratification criteria developed from clinical and ultrasound information from all patients enrolled in all completed randomized trials enables identification of patients whose PFOs are likely pathogenic rather than incidental and who on average will benefit, rather than be harmed, by PFO closure^3,4^. A particularly important characteristic is the presence or absence of a physiologic right-to-left shunt of large size. In the prospectively validated PFO-Associated Stroke CAusal Likelihood (PASCAL) classification system, the presence of a large physiologic shunt places a patient in either the “probable” or “possible” relatedness categories, both of which benefit from PFO closure^3^. The randomized trials all characterized right to left shunt size in patent foramen ovale based on quantified transesophageal echocardiography (TEE) bubble studies, most commonly defining a large right-to-left shunt (RLS) as appearance of more than 20 microbubbles in the left atrium within 3 cardiac cycles. For clinicians to accurately apply these evidence-based treatment decision algorithms in clinical practice, it is necessary that routine clinical TEE reports provide measures of shunt size using the same approach as that employed in the randomized trials.

Transcranial Doppler (TCD) ultrasound is an alternative method of detection and quantification of right-to-left shunts. TCD and TEE have been shown to correlate fairly well in determining presence or absence of a PFO,^5^ but their concurrence in determining shunt size has not been well delineated. Formal TCD criteria, the Spencer Grading system, have been established for quantifying RLS size^6^. Similarly to TEE, for clinicians to informatively apply TCD findings in clinical practice, it is necessary that routine clinical TCD reports provide measures of shunt size using the validated formal quantification system.

The primary objective of this study was to delineate the frequency with which RCT-concordant, formal descriptors of RLS size are present in TEE reports in routine clinical practice. Additional objectives were to delineate the frequency with which formal descriptors of RLS size are present in TCD reports in routine clinical practice and the correlation between TEE and TCD measures of shunt size.

## Methods

### Patient selection

We analyzed a prospectively maintained database of consecutive ischemic stroke admissions to the University of California Los Angeles Medical Center, a tertiary care academic center, from October 2022 to September 2023. Inclusion criteria for this study were: 1) imaging-confirmed diagnosis of ischemic stroke; and 2) transesophageal echocardiogram including bubble study performed during the inpatient diagnostic workup. The exclusion criterion was: 1) confirmed non-PFO cardiac or pulmonary source of right-to-left shunting (e.g., tetralogy of Fallot, pulmonary arteriovenous malformation).

### Data Collection

This study was approved by the University of California Los Angeles Institutional Review Board and performed under a waiver of explicit consent for a minimal risk study of existing electronic medical record data.

Patient demographic and clinical characteristics analyzed included age, sex, presenting stroke severity [assessed with the National Institutes of Health Stroke Scale (NIHSS) score], and ischemic stroke etiologic subtype as classified by the Trial of Org 10172 in Acute Stroke Treatment (TOAST) criteria.^7^

### Transesophageal Echocardiogram Clinical Reports and Formal Re-Analysis

Transesophageal echocardiograms were conducted and interpreted by board-certified cardiologists. For the analysis of clinical TEE findings, the following data were abstracted from TEE written reports entered into the electronic medical record: 1) method of assessment for PFO (bubble study, Doppler study, none); 2) if bubble study performed, whether the report stated that the number of crossing microbubbles was formally quantified; 3) descriptive terms employed to characterize shunt size; 4) whether microbubbles appeared in the early or late phase after injection.

Reports stating only non-quantified descriptive terms for shunt size were bundled into three broad groupings: a) Small (indicated by terms such “small”, “mild”, or “trace”; b) Moderate (indicated by terms such as “moderate”, “medium”, or “intermediate”); and c) Large (indicated by terms such as “large”, “substantial”, “very large”). For analyses applying these groupings within the PFO-Associated Stroke Causal Likelihood (PASCAL) Classification system, which recognizes a large PFO as an indication for PFO closure, a small PFO was considered not a strong indication for closure, a moderate PFO was considered to be an uncertain indication for closure, and a large PFO was considered to be a strong indication for closure.

In each case in which quantified shunt size measurements were made, the reporting cardiologist indicated the following scale was employed, based on the counted number of microbubbles in the left atrium within the three cardiac cycles from bubble infusion: Very small (1 to 9 microbubbles), Small (grade 2, 10-20 microbubbles), and Large (grade 3, >20 microbubbles). However, this scale is an adaptation of a scale developed for rating shunt size on transthoracic, not transesophageal, echocardiograms^8^. Therefore for analyses within the PASCAL classification system, this scale was remapped to the dichotomous grouping employed in the PFO closure RCTS: Small (1-20 microbubbles) – not a strong indication for closure; and Large (>20 microbubbles) – a strong indication for closure.

Formal re-analysis of shunt size on all TEE bubble studies demonstrating a PFO was performed by a study core lab panel comprising two board-certified cardiologists, one with expertise in echocardiography (SD) and one with expertise in PFO management (JT). The cardiologists jointly re-reviewed TEE images saved on Syngo^TM^. PFO size class on formal re-review was assigned based on the counted number of microbubbles in the left atrium within the three cardiac cycles from bubble infusion: Small (grade 1, 1 to 9 microbubbles), Moderate (grade 2, 10-20 microbubbles), and Large (grade 3, >20 microbubbles).

### Transcranial Doppler RLS Estimation

Only TCD studies conducted within 1 month of post-stroke TEE performance were included to minimize confounding by intercurrent events. All TCD bubble studies followed the institutional protocol of infusion of agitated saline mixed with 1ml blood into an antecubital vein and detection of high-intensity transient signals using M-mode technology, along the direction of blood flow in bilateral middle cerebral arteries for 1 minute. Patients were asked not to perform a Valsalva maneuver on the first bubble study run, and the bubble studies were repeated twice with a Valsalva exertion immediately prior to the infusion, followed by relaxation for 10 seconds to augment RLS. Shunt size was then characterized on all clinical reports using the Spencer rating system: grade 0 [0 high intensity transient signals (HITS), no evidence of RLS], grade 1 (1-9 HITS, miniscule RLS), grade 2 (11-30 HITS, minimal RLS), grade 3 (31- 100 HITS, mild RLS), grade 4 (101-300 HITS, moderate RLS), grade 5 (>300 or a shower of HITS, large RLS)^6^. The clinical TCD report was interpreted by a vascular neurology fellow and a supervising board-certified vascular neurologist. For analyses in the current study comparing TCD ratings to the trichotomous TEE ratings, this scale was remapped to a three-level scale: Small (grade 1/2), Intermediate (grade 3), and large (grade 4/5). For analyses applying these groupings within the PASCAL Classification system, a small PFO was considered to be not a strong indication for closure, an intermediate PFO was considered to be an uncertain indication for closure, and a large PFO was considered to be a strong indication for closure.

### Statistical analysis

Descriptive statistics were used to characterize the frequency of PFO presence per clinical TEE reports, in all patients and separately in patients with TOAST diagnoses of known cause or undetermined cause of stroke. Descriptive statistics were used to characterize the frequency of PFO presence per clinical TCD reports and degree of agreement with TEE regarding presence/absence of PFO. The primary outcomes were the frequency with which formal, clinical-trial adherent characterization of PFO shunt size descriptors were used, and the proportion of concordant size estimation between TEE and TCD.

Statistical comparisons were performed between the patients with detected RLS and those without from their TEE bubble study. Variables are recorded as means with standard deviations or medians with interquartile range (IQR) for continuous variables (Mann Whitney U tests for comparison). Categorical variables were reported as numerical counts, with comparisons performed by Pearson Chi-squared analysis if cell sizes were all 5 or more. If any cell size was less than 5, then Fisher’s exact test was used instead.

In the TEE-TEE analysis, concordance in RLS shunt size categories was assessed between TEE clinical reports and the TEE quantitative re-review. In the TEE-TCD analysis, concordance in RLS shunt size categories was assessed between TEE clinical reports and TCD clinical reports (which included quantification in all instances). For the TEE-TCD comparison, the pattern of congruence and noncongruence was delineated using a concordance bubble chart.

We followed the Strengthening the Reporting of Observational Studies in Epidemiology (STROBE) reporting guideline^9^.

## Results

Overall, 77 ischemic stroke patients underwent same-admission inpatient TEE during the study period, among whom shunt presence was assessed by bubble study in 60 (77.9%), direct color Doppler alone in 5 (6.5%), and not assessed in 12 (15.6%). Of those 17 who did not undergo a bubble study for the presenting stroke, 5 patients had no intracardiac shunt according to direct color Doppler, 5 patients had prior bubble studies already showing no shunt, 1 patient had a prior study at an outside facility already reported to demonstrate a shunt, 2 patients had the TEE performed to evaluate for atrial thrombi prior to cardioversion of atrial fibrillation, and 9 patients did not have a documented reason.

Among the 60 patients with TEE bubble study, right-to-left shunt was deemed present on TEE in 25 (42%) and absent in 35 (58%). Table 1 shows baseline patient characteristics of patients with and without RLS on TEE. Patients with RLS had a significantly lower frequency of tobacco use and non-significantly higher stroke deficit severity scores on the National Institutes of Health Stroke Scale.

**Table 1:**
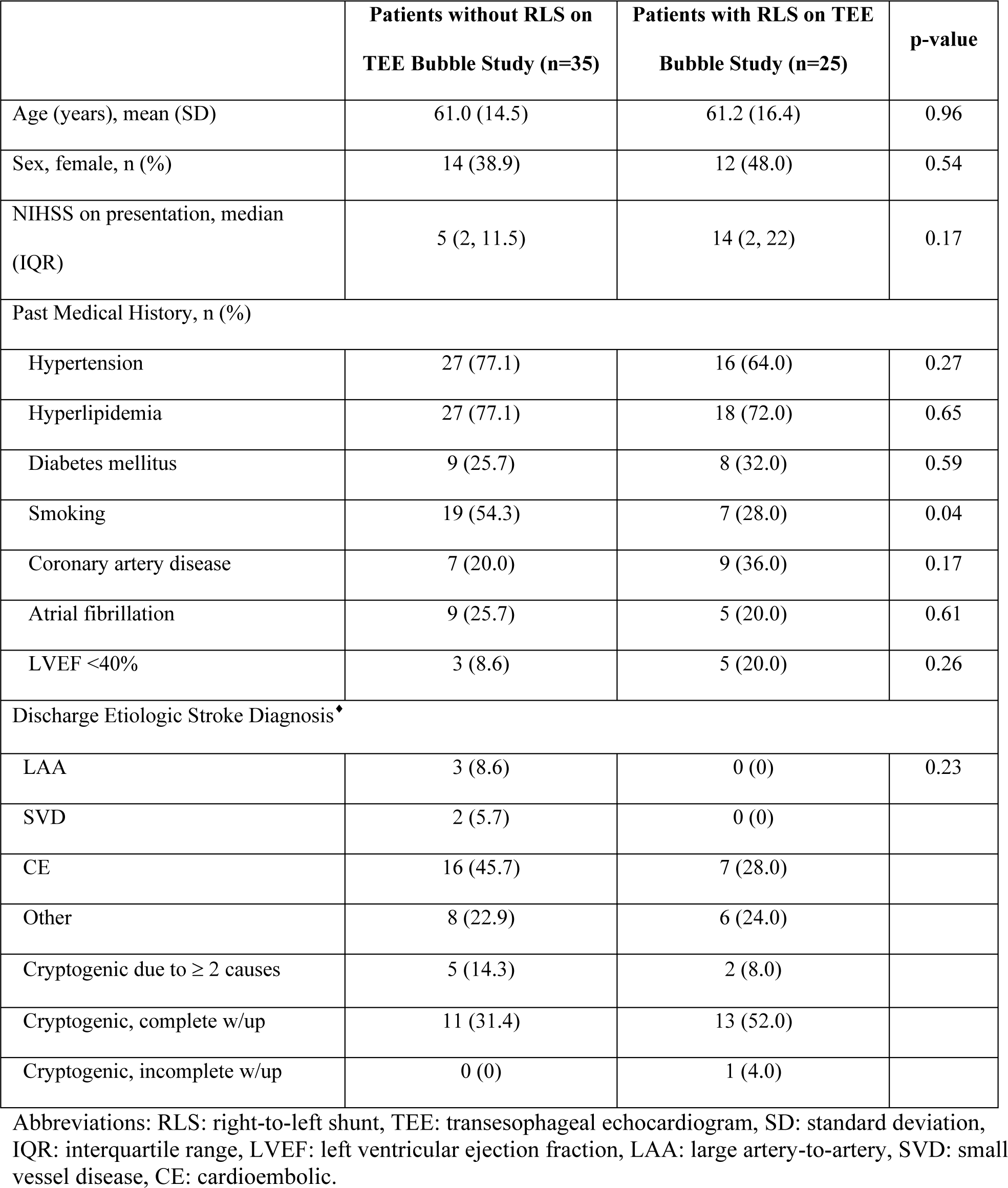
Baseline Characteristics and Discharge Etiologic Stroke Diagnosis in Patients without vs. with detected right-to-left shunt on TEE Bubble Study.

Among the 25 patients with RLS on TEE, the TEE report in 21 (84%) patients only informally characterized shunt severity with various descriptive terms without providing quantitative data. A total of six different descriptive terms were used, with “small/mild/trace” in 13 cases, “moderate/medium” in 6, and “large” in 1. In one other patient, bubbles appeared in the left atrium after an extended delay, and the report indicated that a non-PFO RLS was present with shunt size not characterized either formally or informally. Clinical reports formally quantified shunt size in 4 (16%) patients, rating shunt size as small (1-9 bubbles) in one and medium (10-19 bubbles) in three.

TCD bubble studies were performed in 19/25 (76%) of the patients who had evidence of RLS on TEE. In all (100%) TCD reports, shunt size was formally quantified with both exact bubble counts and Spenser classification grades. Among the 19 patients undergoing both TCD and TEE, 79% (15/19) had informal shunt size grading on TEE, 16% (3/19) had formal shunt size grading on TEE, and 5% (1/19) had no shunt size characterization on TEE.

Among the 15 patients with only informal verbal descriptions of RLS size on TEE, the relation of the informal TEE categorizations to formal TCD quantified shunt size categorization is shown in **Figure 1**. Concurrence between TCD and TEE for shunt size across all three size levels (small, intermediate, large) was low, 3/15 (20%). Shunt sizes were categorized as larger on TCD than TEE in 7/15 (47%) and smaller on TCD than TEE in 5/15 (33%). The incongruent size classifications affected presence of the shunt size indication for or against PFO closure in 33% (5/15) patients. In 27% (4/15), TCD identified a large shunt size supporting closure while TEE identified a small shunt size not supporting closure. Conversely, in 7% (1/15), TCD identified a small shunt size not supporting closure while TEE identified a large shunt size supporting closure.

**Figure 1:**
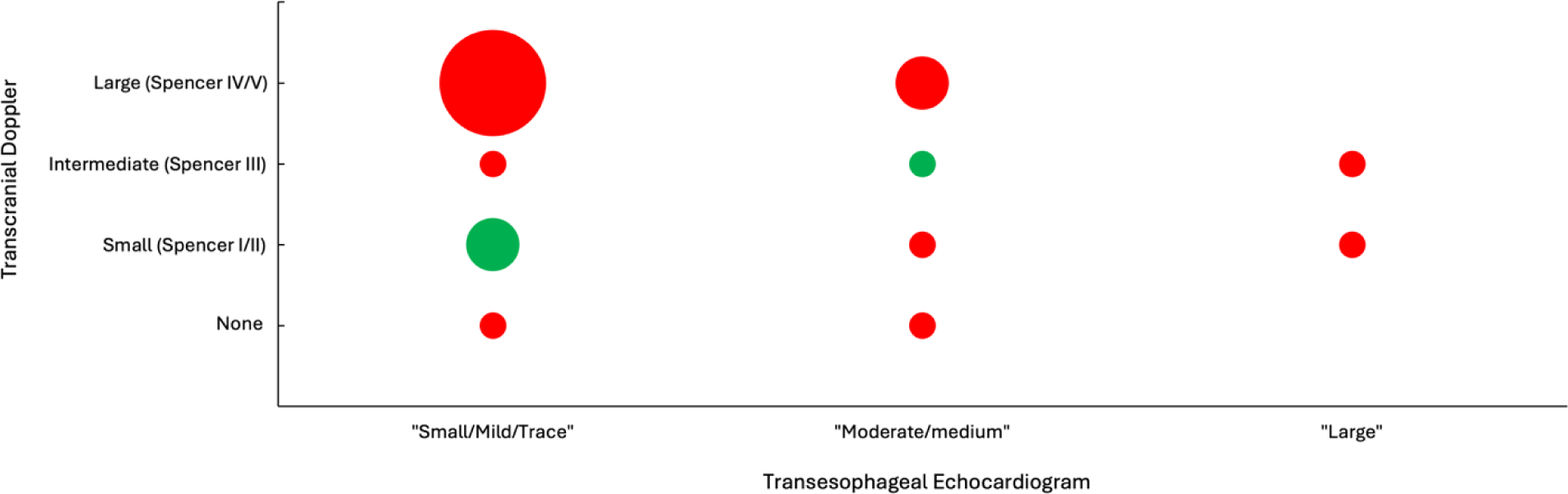
Bubble plot showing agreement of right-to-left shunt size assessments between transcranial Doppler and transesophageal echocardiography clinical reports. Green: concurrence; red: non- concurrence. Bubble size indicates number of cases in each cell.

Among the 3 patients with clinical TEE reports that quantified RLS size who also underwent TCD, both modalities classified shunt size as small or very small in all 3 cases.

The formal quantitative re-scoring of the TEEs in which clinical reports only had verbal descriptions, changed the size classification of 6/15 (40%) patients compared with the original clinical TEE report, with larger size assigned in three and smaller size assigned three **(Figure 2)**. Concurrence across all three shunt size levels (small, intermediate, large) between echo Core Lab quantified TEE re- readings and TCD remained low, with concurrence in 27% (4/15) of patients **(Figure 3)**.

**Figure 2:**
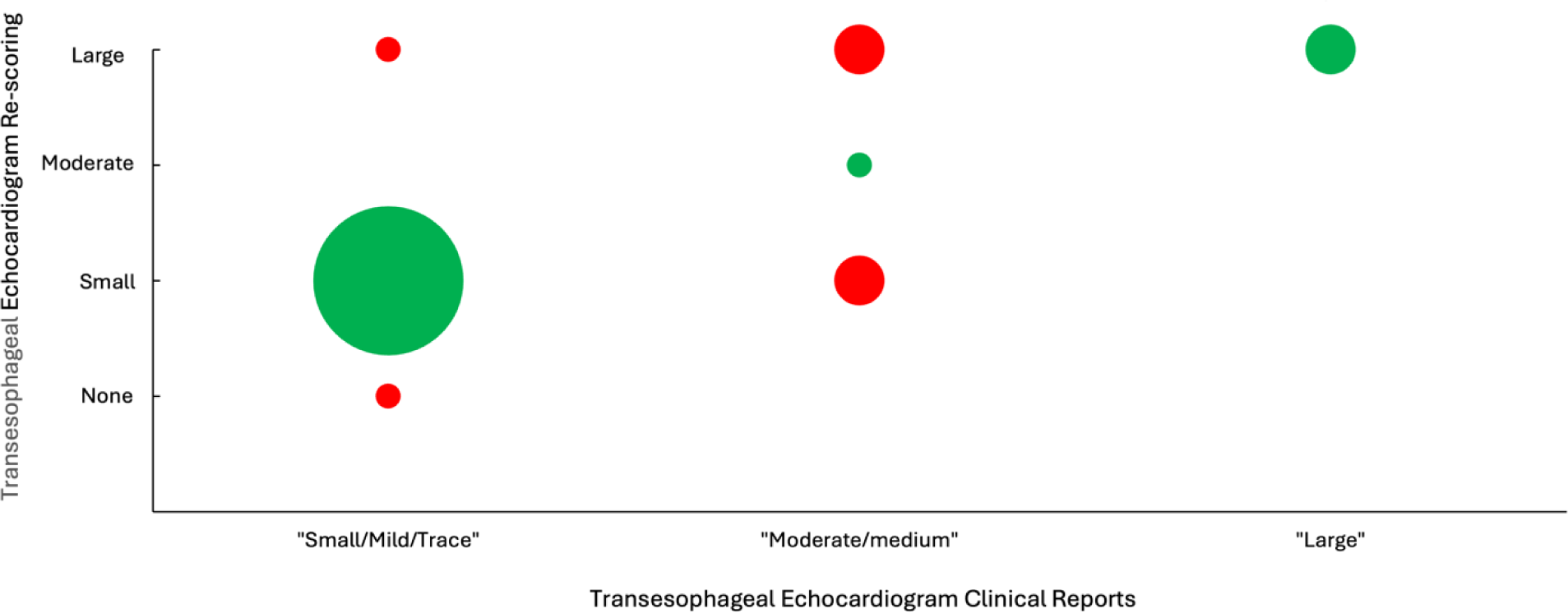
Bubble plot showing agreement of right-to-left shunt size assessments between TEE clinical reports and TEE Core lab. Green: concurrence; red: non-concurrence. Bubble size indicates number of cases in each cell.

**Figure 3.**
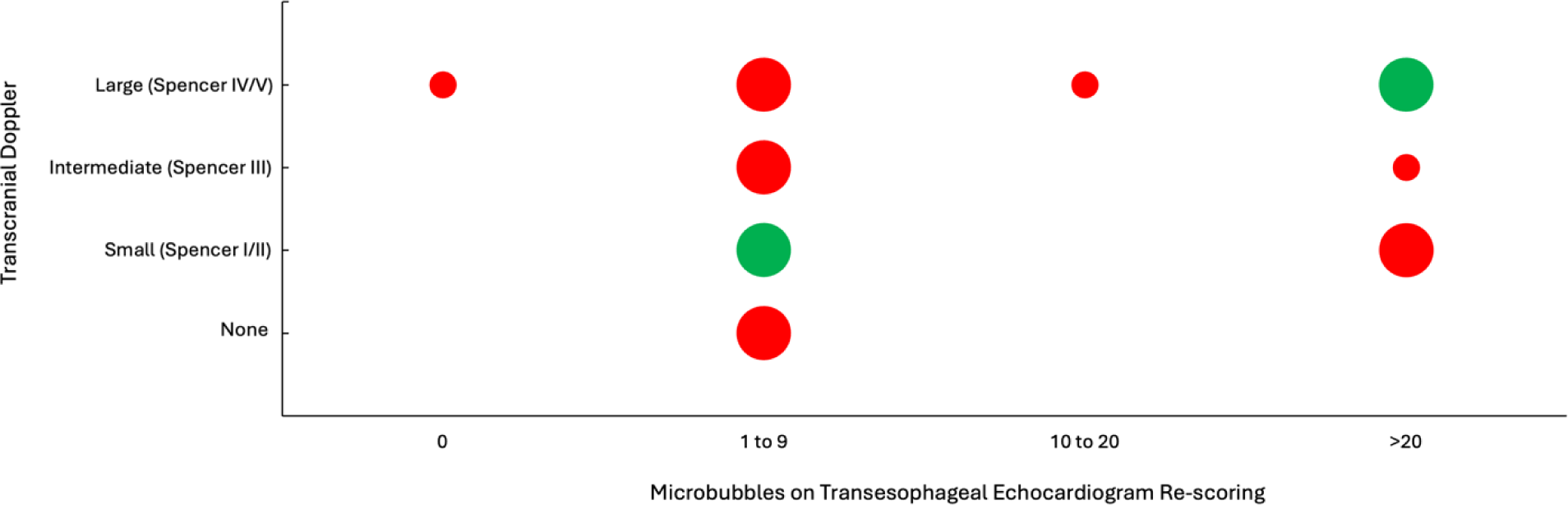
Bubble plot showing agreement of right-to-left shunt size assessments between transcranial Doppler and TEE Core lab. Green: concurrence; red: non-concurrence. Bubble size indicates number of cases in each cell.

## Discussion

In this cohort study, among consecutive acute ischemic stroke patients with right-to-left shunts undergoing bubble study transesophageal echocardiography, quantitative grading of shunt size based on counting of bubbles in the left atrium was uncommonly provided in clinical study interpretations, present in only 1 of every 6 reports. Moreover, verbal descriptions of shunt size correlated poorly with Core Lab quantitative analysis of the same TEE studies. In contrast, in the few instances in which clinical TEE reports provided quantitative shunt size assessments, correlation with the Core Lab quantitative analysis of the same TEE studies was excellent. Transcranial Doppler ultrasound bubble studies did routinely provide quantitative shunt size ratings in clinical reports. The correlation of clinical TEE with formal quantitative TCD shunt size rating was suboptimal, with incongruence in 4 of every 5 cases, including discrepancies that affected indications for PFO closure in fully one-third of cases.

The failure to use clinical trial-validated interpretive criteria would be a moderate, rather than major, issue If interpreting clinicians’ intuitive, holistic judgments of shunt size were accurate. However, in this study clinician intuitive readings were highly discordant with subsequent formal, quantitative analyses. As a result, in one-third of cases, treating clinicians were provided with TEE shunt size interpretations that would lead to incorrect decision-making between treatment with PFO closure devices or medical therapy. Potential pragmatic capital or workflow mitigating factors would be understandable if performing quantitative analysis required highly specialized equipment or software, or was particularly time-consuming. However, these factors are absent. Quantifying left atrial bubble counts during the first three cardiac cycles requires the exact same hardware and software as informal shunt size estimation and extends interpretation time by only 1-2 minutes per case.

Concern about the accuracy of formal, quantified bubble counts on TEE could potentially contribute to echocardiographer reluctance to employ this approach in clinical practice. TEE does have several technical restrictions that limit bubble count accuracy. These include: 1) a TEE visualizes a tomographic slice, not the entirety, of the left atrium, so some bubbles will inevitably be missed; 2) as bubbles do not necessarily travel linearly across the left atrium, the same bubbles may be present across multiple cardiac cycles, leading to double- or triple-counting; and 3) patients may not be able to perform Valsalva maneuvers to the fullest extent during sedation with a probe in place in the esophagus^10^.

However, test imprecision actually provides more reason to pursue, not avoid, a standardized, quantitative interpretive approach that prevents the addition of interpretative variability to acquisition variability. Most importantly, the quantitative analysis technique has been validated in randomized trials to identify patients who will benefit from PFO closure device placement; the intuitive, holistic analysis technique has not been tested in RCTs.

The use of transcranial Doppler ultrasound to classify RLS size raises an interesting dilemma as there are theoretical reasons to expect TCD to be more accurate than TEE in quantifying shunt size but patient selection using TCD has not been validated in randomized clinical trials. In such circumstances, adding the more accurate TCD test to the clinically validated TEE test is desirable as it will improve identification of the underlying biomarker (large shunt size) responsible for clinical trial findings.

Advantages of TCD in quantifying shunt size include: 1) bubble counts reflect emboli passing through the entire volume of the middle cerebral artery; 2) bubbles are counted individually as they serially traverse the middle cerebral artery, avoiding double- or triple-counting; and 3) patients are able to perform Valsalva maneuvers to the optimal extent. These considerations accord with the finding in the current study that performance of TCD in addition to TEE increased the number of patients identified as having a large shunt who might therefore benefit from PFO closure.

The failure to incorporate clinical trial validated interpretation criteria into clinical TEE reports documented in this study is a cardinal example of the well-known gap between research evidence and clinical practice. Across all medical disciplines, it takes an average of 17 years for new knowledge generated by randomized clinical trials to be incorporated into practice, and even then application is highly uneven^11,12^. A systematic review identified seven barriers to translating evidence into practice: cognitive-behavioral barriers, attitudinal or rational-emotional barriers, professional barriers, barriers embedded in guidelines or evidence, patient barriers, support or resource barriers, and system and process barriers^13^. While all of these are likely contributory in this instance, especially notable is the barrier consisting of the absence of guideline endorsement of reporting quantified classification of shunt size on clinical TCD reports. This barrier is readily correctible and likely to have a major impact upon care^14,15^.

This study has limitations. First, the study was performed at a single academic medical center, albeit an accredited Comprehensive Stroke Center with the highest quality achievement levels and an echocardiography lab accredited by the Intersocietal Accreditation Commission for the Accreditation of Echocardiography Laboratories. Assessment in a greater number and wider range of hospitals is desirable. Second, no reference standard was available to provide ground truth for ratings of RLS shunt size. The clinical gold standard of the presence of a PFO would be passage of a guidewire across the atrial septum during a right heart catheterization. Third, the sample size was moderate, limiting study power.

Replication in large studies would be worthwhile.

## Conclusions

Quantified, evidence-based ratings of PFO shunt severity were present in only 1 of every 6 TEE reports, and unquantified, informal size estimates correlated poorly with TCD quantification of shunt severity. Patient management would be aided by inclusion of formal PFO shunt size quantification in all clinical stroke patient TEE reports.

## Data Availability

The raw data supporting the conclusions of this article will be made available by the authors upon reasonable requests, without undue reservation.

## Funding Sources

None

## Disclosure Statement

P.Y.Sun: None. J.M.Tobis: n/a. D.S.Liebeskind: Consultant; ; Cerenovus, Genentech, Medtronic, Stryker, Rapid Medical. R.C.Alfonso: None. J.L.Saver: Consultant; ; Medtronic, Abbott, Roche, BrainsGate, BrainQ, Bayer, CSL Behring, Rapid Medical, Novo Nordisk, Individual Stocks/Stock Options; ; QuantalX.

